# Differences by country-level income in COVID-19 cases, deaths, case-fatality rates, and rates per million population in the first five months of the pandemic

**DOI:** 10.1101/2020.07.13.20153064

**Authors:** Wrihsmeen Sabawoon

## Abstract

**Objective:** To describe differences by country-level income in COVID-19 cases, deaths, case-fatality rates, incidence rates, and death rates per million population.

**Methods:** Publicly available data on COVID-19 cases and deaths from December 31, 2019 to June 3, 2020 were analyzed. Kruskal-Wallis tests were used to examine associations of country-level income with COVID-19 cases, deaths, case-fatality rates, incidence rates, and death rates.

**Results:** A total of 380,803 deaths out of 6,348,204 COVID-19 cases were reported from 210 countries and territories globally in the period under study, and the global case-fatality rate was 6.0%. Of the total globally reported cases and deaths, the percentages of cases and deaths were 59.9% and 75.0% for high-income countries, and 30.9% and 20.7% for upper-middle-income countries. Countries in higher income categories had higher incidence rates and death rates. Between April and May, the incidence rates in higher-income groups of countries decreased, but in other groups it increased.

**Conclusions:** In the first five months of the COVID-19 pandemic, most cases and deaths were reported from high-income and upper-middle-income countries, and those countries had higher incidence rates and death rates per million population than did lower-middle and low-income countries.

## Introduction

The disease caused by a coronavirus that recently emerged in Wuhan, China (He et al., 2020) is now known as COVID-19. Most patients with COVID-19 have a respiratory illness with flu-like symptoms including cough, fever, fatigue, and shortness of breath. Some have pneumonia, acute respiratory distress syndrome, respiratory failure, and multiple organ failure, and a few die (Huang et al., 2020). The causative agent is a previously unknown coronavirus: Severe Acute Respiratory Syndrome (SARS)-CoV-2 (Alexander et al. 2020; Andersen et al. 2020; Lu et al. 2020; WHO 2020a; WHO 2020b; Zhou et al. 2020; Zhu et al. 2020).

The COVID-19 outbreak started a few weeks before the Chinese Spring Festival, which usually occurs in January or February on the Gregorian calendar. In 2020 it was celebrated January 24-30. Each year around the time of that festival, many Chinese people visit their ancestral homes and some people from outside China visit the country for sightseeing and to celebrate the start of the lunar year. In 2020, that seasonal increase in travel exacerbated the spread of SARS-CoV-2 within China and also to countries and territories outside China. The worldwide pandemic was recognized as such on March 11, 2020, after more than 118,000 cases had been reported in 110 countries and there was judged to be a sustained risk of further global spread (Bedford et al. 2020). As of mid-March, 146 countries were affected, 154,000 people had been infected, and 5,700 fatalities had been recorded. In absolute terms, the number of cases was very high in China (81,048 cases), followed by Italy (21,157), Iran (12,729), the Republic of Korea (8162), Spain (5753), France (4469), Germany (3795), the US (1678), Switzerland (1359), and the UK (1144). On March 23, 2020, the WHO reported that little had changed in China (81,601 cases), but Italy had already recorded 59,138 cases, Spain 28,572, Germany 24,774, and France 15,821 (Welfens 2020). By the end of March 2020, COVID-19 cases had occurred in all 50 states of the US. At the time of this writing (May 24, 2020), 210 countries and territories had reported cases of SARS-CoV-2 infection, and the US had the most confirmed active cases and deaths in the world (Data from Our World in Data).

The numbers of cases and deaths from COVID-19 differ both among countries (Johns Hopkins University data) and within countries (CDC COVID-19 response team 2020). These differences might be caused by differences in the timing and early transmission of the virus, by countries’ response patterns, by the numbers of tests conducted, by sociodemographic conditions, by connectivity and trade patterns, and by environmental factors. Pandemics generally have their greatest effects on lower-middle-income and low-income countries and on socially disadvantaged populations, because they intensify existing health inequalities. During the 1918 “Spanish” influenza pandemic, racial minorities had higher all-cause mortality and influenza mortality rates than did Caucasians (Hutchins et al. 2009). In the 2009 H1N1 influenza pandemic, minority groups had higher rates of serious infection requiring hospitalizations than did non-minority groups (CDC 2020). Nonetheless, so far during the COVID-19 pandemic, more cases and deaths have been reported from high-income and upper-middle income countries. In the absence of a vaccine, given that responses differ between countries, and given the fact that information regarding transmission is still insufficient, it is difficult to confidently predict the course of the pandemic as it applies to individual countries. The virus has also reached almost all low-income and middle-income countries, where community transmission has started. Most of those countries have also closed their borders to prevent travel-related dissemination of the virus. Those include countries with large populations living in overcrowded conditions where “social distancing” is impossible to maintain, and with health systems that are unlikely to be able to meet the challenges of this pandemic.

In some previous studies, data on COVID-19 cases and deaths were analyzed to detect and predict seasonality of the disease (Bukhari and Jameel 2020; Sajadi et al. 2020) and to describe cases and deaths by continent and by country (Lai et al. 2020). To the best of our knowledge, no previous study has described the distribution of cases and deaths by country-level income. Such an analysis has analytical and practical purposes. Looking at countries as members of income-defined groups can highlight trans-national phenomena regarding how economic activity is associated with the spread of the disease and with the resulting burdens on healthcare systems. The findings from such analysis can also be used by policy makers, international organizations, and bilateral agencies to allocate resources for the control of the epidemic according to income-defined groups. We therefore describe the worldwide distribution of the numbers of cases and deaths by country and by country-level income status; describe trends in the numbers of cases and deaths; and examine how country-level income status is associated with COVID-19 case-fatality rate and with the numbers of cases and deaths per million population in the first five months of the pandemic. For that purpose we consider each country’s income status, using the World Bank’s approach, classifying countries into high-income, upper-middle-income, lower-middle-income, and low-income groups (Fantom and Umar 2016).

## Methods

Two sources of publicly available data were used. First, data on COVID-19 cases and deaths in all affected countries were downloaded from the Our World in Data (OWID) website. The OWID’s data on cases and deaths come from the European Centre for Disease Prevention and Control; data on testing were collected from formal reports of countries by the data team of OWID; data on other variables came from the United Nations, the World Bank, the Global Burden of Disease Collaborative Network, global burden of disease study 2017 results, the Blavatnik School of Government, the Oxford COVID-19 Government Response Tracker, and national governmental reports. The data are updated daily, and they contain the dates of daily reported data, the country or territory name, the numbers of total and new cases and deaths, the numbers of total and new cases and deaths per million population, and the numbers of total and new tests and their rates per 1,000 population, among other variables (OWID data 2020^a^).

Second, data classifying countries by their income status were downloaded from the World Bank website and incorporated into the master dataset. The countries were classified according to the World Bank classification of countries by income, which has been used since 1989. It divides countries into four groups — low income, lower-middle income, upper-middle income, and high income — using gross national income per capita valued annually in US dollars using a three-year average exchange rate. The classification is published at http://data.worldbank.org and revised versions are published each year on July 1 (Fantom and Umar 2016). The World Bank did not provide data on income for seven small areas, all of which reported cases of COVID-19: Jersey, Guernsey, the Falkland Islands, Vatican City State, Montserrat, Bonaire Sint Eus, and Anguilla. Therefore, those seven were categorized as “small areas with unknown income status”. The data were exported to STATA 2012 for analysis.

Cases on international conveyances (e.g. cruise ships) were excluded from the analysis, because the patients were from many countries, and therefore as groups they could not be linked with any of the World Bank’s income categories.

Descriptive statistics regarding cases, deaths, case-fatality rates (as percentages), cases per million population, and deaths per million population for each country and for income-defined groups of countries were calculated. Kruskal-Wallis one-way analysis of variance on ranks was used to examine associations of country-level income category with the variables listed above. Data from the small areas with unknown income status were excluded from the Kruskal-Wallis analysis because there was no information on country-level income status, and because the numbers of cases were much smaller than in the other income-groups. One-way analysis of variance (ANOVA) was used to examine associations of country-level income category with the time since the onset of the pandemic.

Changes over time in the indices mentioned above were also examined for trends during the period for which data were available.

## Results

### Expansion period of the pandemic

The first cases of COVID-19 were reported to the WHO on December 31, 2019. By June 3^rd^, 2020, the disease had spread to 210 countries and territories (Table 1). The mean number of days from the time that the first case was reported to the WHO until June 3, 2020 differed widely among income-defined groups: 100.5 (SD 19.6) days for high-income countries; 94.8 (SD 18.6) for upper-middle-income countries; 89.5 (SD 20.1) for lower-middle-income countries; and 79.1 (SD 17.2) for low-income countries (F= 870.36, df = 3, p < 0.001).

**Table 1:**
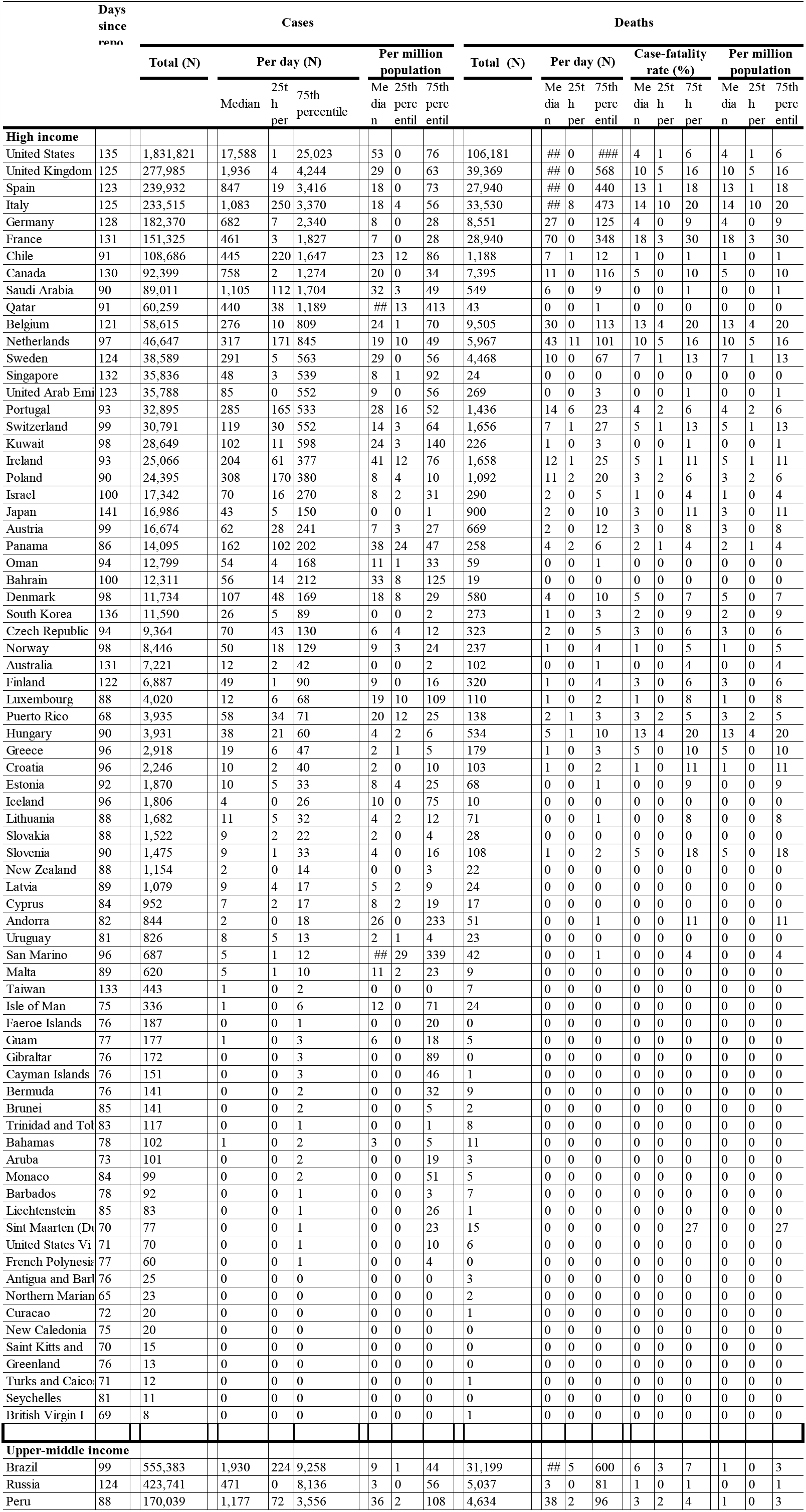
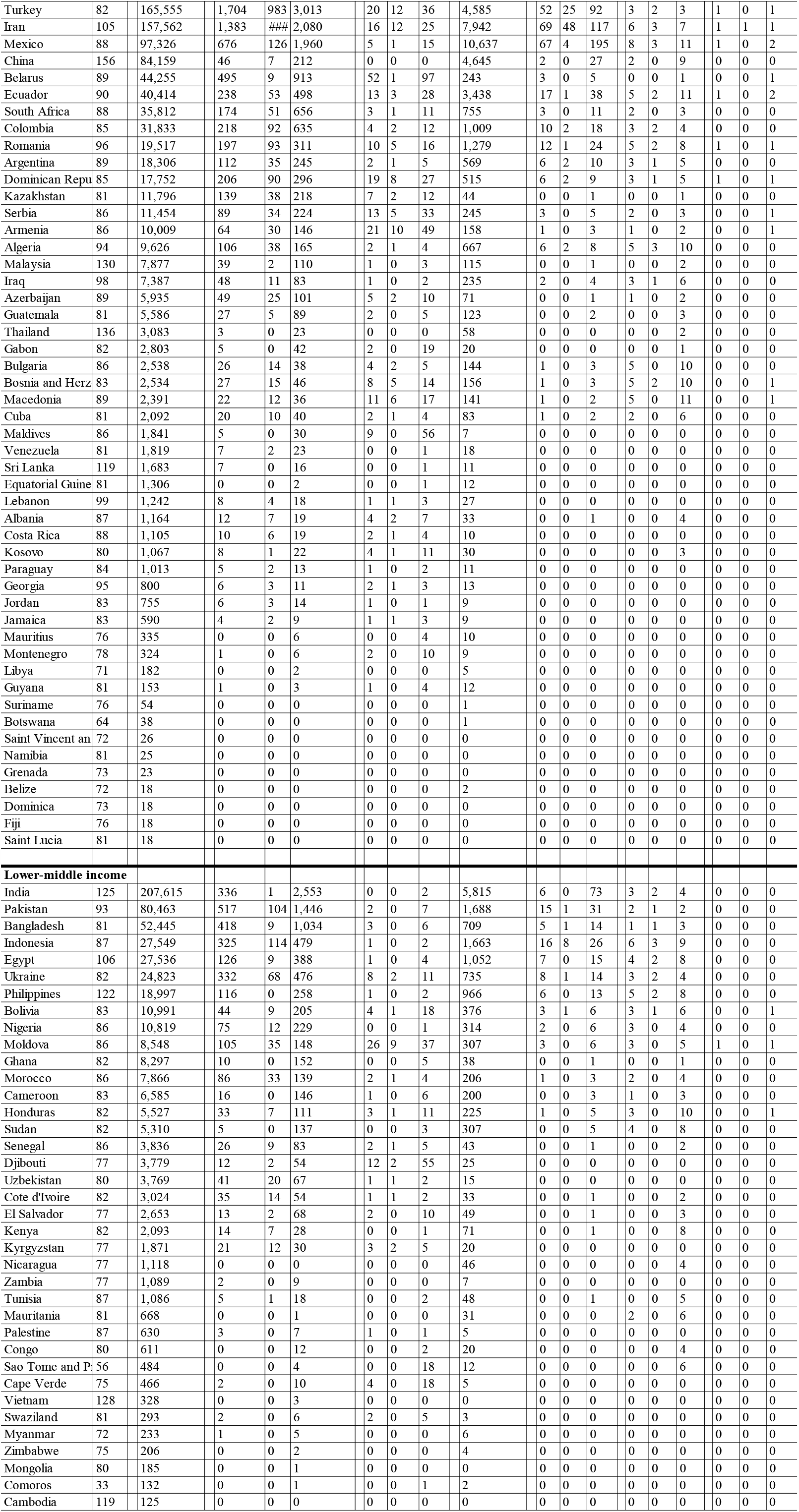
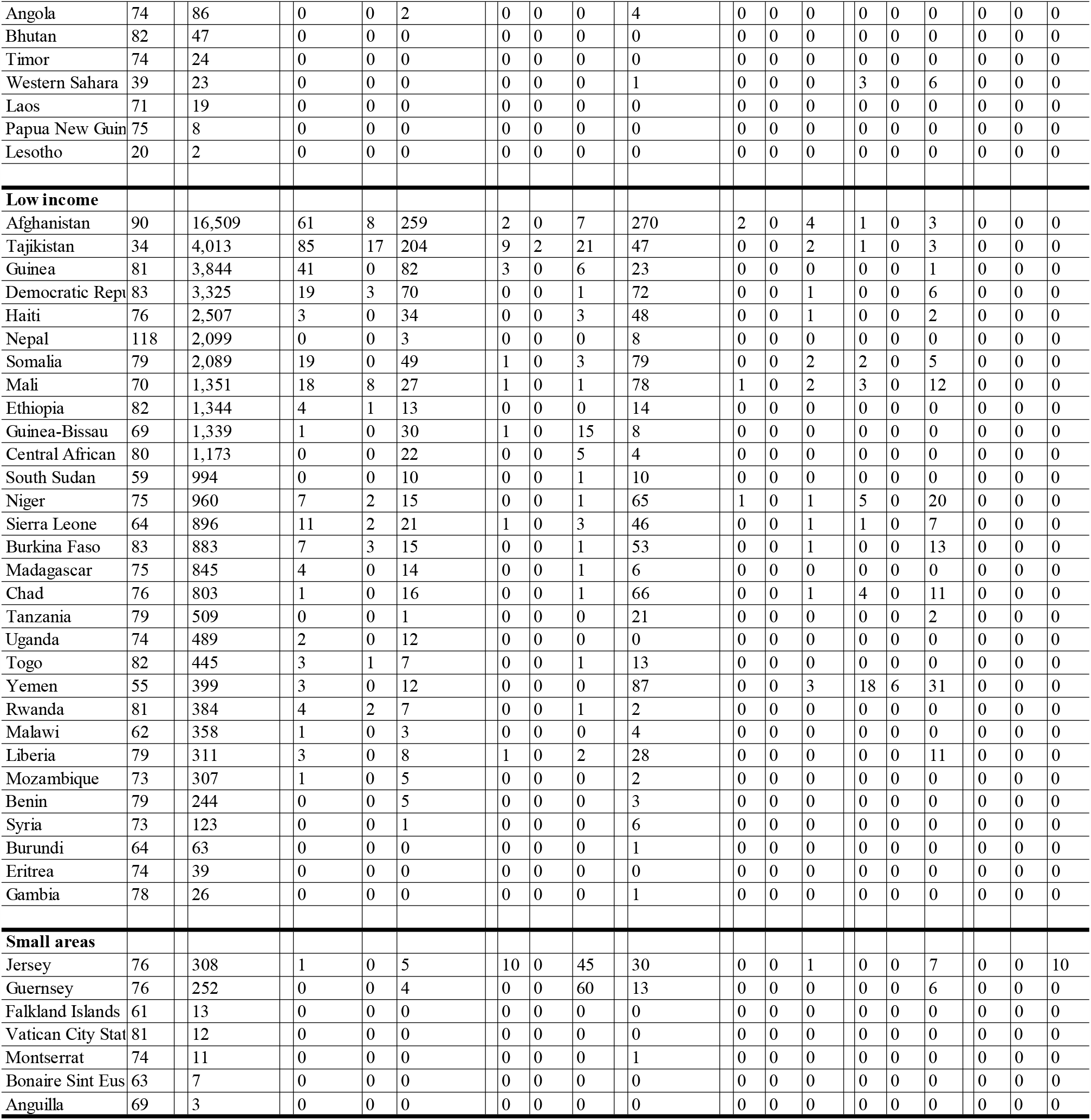
Numbers of COVID-19 total and daily cases and deaths; their rates per million population, and case-fatality rates from the onset of the pandemic until June 3rd, 2020, by country.

### Numbers of reported cases and deaths

In five months, 6,804,286 cases were reported globally. About 90% were reported from high-income or upper-middle-income countries (Table 2). The number of reported cases within each income-defined group varied. The top-10 high-income countries accounted for 48.0% of all COVID-19 cases globally. The top-10 upper-middle-income countries accounted for 26.1% of the cases globally. In contrast, the top-10 lower-middle-income countries accounted for only 6.9%, and the top-10 low-income countries accounted for only 0.6%. The remaining 170 countries and territories reported only 18.4% of all COVID 19 cases globally (Table 1). Countries in higher income categories reported more cases per day (*χ*^2^ = 514.79, df = 3, p < 0.001) (Table 2).

**Table 2:** Contribution of countries by income group to global numbers of COVID-19 cases and deaths and their daily reported cases, deaths, and case-fatality ratios from the onset of the pandemic until June 3rd, 2020.

| Variable | Income category | Total | % Global | Daily reported data |  |  |
| --- | --- | --- | --- | --- | --- | --- |
|  |  |  |  | Median | 25th percentile | 75th percentile |
| Total cases |  |  |  |  |  | p<0.001 |
|  | High | 3,804,286 | 59.9 | 10.0 | 0.0 | 147.0 |
|  | Upper-Middle | 1,962,382 | 30.9 | 14.0 | 1.0 | 111.0 |
|  | Lower-Middle | 532,259 | 8.4 | 5.0 | 0.0 | 67.0 |
|  | Low | 48,671 | 0.8 | 2.0 | 0.0 | 15.0 |
|  | Small areas with unknown income status | 606 | 0.0 | Excluded |  |  |
|  | Global | 6,348,204 | 100.0 | 7.0 | 0.0 | 75.0 |
| Total deaths |  |  |  |  |  | p < 0.001 |
|  | High | 285,666 | 75.0 | 0.0 | 0.0 | 3.0 |
|  | Upper-Middle | 78,977 | 20.7 | 0.0 | 0.0 | 3.0 |
|  | Lower-Middle | 15,051 | 4.0 | 0.0 | 0.0 | 1.0 |
|  | Low | 1,065 | 0.3 | 0.0 | 0.0 | 0.0 |
|  | Small areas with unknown income status | 44 | 0.0 | Excluded |  |  |
|  | Global | 380,803 | 100.0 | 0.0 | 0.0 | 2.0 |
| Case-fatality rate |  |  |  |  |  | p < 0.001 |
|  | High | NA | NA | 0.6 | 0.0 | 7.0 |
|  | Upper-Middle | NA | NA | 0.7 | 0.0 | 4.2 |
|  | Lower-Middle | NA | NA | 0.0 | 0.0 | 3.4 |
|  | Low | NA | NA | 0.0 | 0.0 | 2.3 |
|  | Small areas with unknown income status | NA | NA | Excluded |  |  |
|  | All of the above | NA | NA | 0.0 | 0.0 | 4.8 |

**P-value for Kruskal-Wallis test (small areas excluded).**

The high-income and upper-middle-income countries also accounted for most of the reported deaths. In five months, 380,803 deaths were reported, and thus the global case-fatality rate was 6.0%. Three quarters of all deaths were reported from high-income countries and one fifth were reported from upper-middle-income countries. Countries in higher income categories reported more deaths per day (*χ*^2^= 349.92, df = 3, p < 0.001) (Table 2).

### COVID-19 incidence and mortality rates per million population

The overall median daily incidence rate of COVID-19 for all affected countries in the period was 0.9 per million population (interquartile range: 0.0 to 9.3). The incidence rate of the disease varied across countries (Table 1), and it was higher in countries with higher incomes (*χ*^2^ = 1440.98, df = 3, p < 0.001) (Table 3). The overall median daily death rate per million population was 0.0 (interquartile range: 0.00 to 0.01), which also differed between income-defined groups of countries. The 75^th^ percentile of the death rates for high-income, upper-middle-income, lower-middle-income, and low-income groups were 0.5, 0.2, 0.0, and 0.0, respectively (*χ*^2^ = 479, df = 3, p < 0.001) (Table 3).

**Table 3:** Association of country-level category with COVID-19 incidence and death rates per million population from the onset of the pandemic until June 3rd, 2020.

| Variable | Income category | Median | 25th percentile | 75th percentile |
| --- | --- | --- | --- | --- |
| **Cases per million population** |  |  |  | p < 0.001 |
|  | High | 4.2 | 0.0 | 26.2 |
|  | Upper-Middle | 1.5 | 0.0 | 8.6 |
|  | Lower-Middle | 0.3 | 0.0 | 2.6 |
|  | Low | 0.1 | 0.0 | 0.8 |
|  | Small areas with unknown income status | Excluded |  |  |
|  | All above | 0.9 | 0.0 | 9.3 |
| **Deaths per million population** |  |  |  | p < 0.001 |
|  | High | 0.0 | 0.0 | 0.5 |
|  | Upper-Middle | 0.0 | 0.0 | 0.2 |
|  | Lower-Middle | 0.0 | 0.0 | 0.0 |
|  | Low | 0.0 | 0.0 | 0.0 |
|  | Small areas with unknown income status | Excluded |  |  |
|  | All above | 0.0 | 0.0 | 0.1 |

**P-value for Kruskal-Wallis test (small areas excluded).**

### Trend in the numbers of cases, deaths, and their rates per million population

From the onset of the pandemic until June 3, 2020, the total number of cases increased globally. In the high-income countries, of the total 3,804,286 reported cases, 0.1%, 16.2%, 45.9%, and 34.8% were reported in February, March, April, and May, respectively. A decline in cases was recorded between April and May. In the upper-middle-income group, over the same four months the number of cases increased. Of the total reported cases (1,962,382), 3.6%, 4.2%, 24.4%, and 60.1% were reported in February, March, April, and May, respectively. In the-lower middle-income group, that month-over-month increase in the percentage of cases increased substantially in May. Of the total cases in this income group (532,259), 2.0% were reported in February and 67.1% in May. In low-income countries, the percentage of cases increased between February and May from 0.0% to 71.1%. Similarly, the median (interquartile range) of the daily incidence rates per million population in high-income countries declined between April and May. It was 0.0 (0.0 to 0.1), 5.6 (0.8 to 23.3), 13.0 (0.3 to 45.0), and 2.9 (0.0 to 20.9) per million population in February, March, April, and May, respectively. However, during the same time period, the daily incidence rates per million population increased in all other groups: from 0.0 (0.0 to 0.1) to 2.9 (0.0 to 14.0) in upper-middle-income countries, from 0.0 (0.0 to 0.0) to 1.5 (0.0 to 6.0) in lower-middle-income countries, and from 0.0 (0.0 to 0.0) to 0.4 (0.0 to 1.9) in low-income countries.

In high-income countries the number of deaths also declined between April and May. Among all deaths (285,666), 0.0% were reported in February. That rose to 57.9% in April but then declined to 29.5% in May. In contrast, between February and May the number of cases in upper-middle-income, lower-middle-income, and low-income countries increased. Among all deaths in the upper-middle-income group (78,977), 3.4% were reported in February, 25.1% in March, and 60.1% in May. Among all deaths in the lower-middle-income group (15,051), 0.0% were reported in February, 25.7% in March, and 61.8% in May. Among all deaths in the low-income group (1,065), 0.0% were reported in February, 24.3% in April, and 66.5% in May. Similarly, the death rate per million population declined in high-income countries and was approximately constant in the remaining three income groups.

In the small areas with no income data, both cases and deaths declined after April.

## Discussion

As of June 3, 2020, a total of 6,804,286 COVID-19 cases including 380,803 (6.0%) deaths had been reported globally. Income-defined groups of countries differed greatly in the numbers of cases and deaths and in their rates, as follows.

### Income-related differences in cases and in incidence rates per million population

Among the main findings of this study are the income-related differences in COVID-19 cases and their incidence rates per million population. The income-defined category of a country was positively associated with the number of COVID-19 cases and with the daily incidence rate per million population. The large differences in daily incidence rates probably reflect differences in epidemiologic and population characteristics, and differences in clinical and public health practices.

First, the most obvious of the epidemiological differences is in the timing of the introduction and early transmission of SARS-CoV-2 in the various countries. Among the income-defined groups, strong connections by air travel probably resulted in the relatively early and widespread transmission of the disease among high-income and upper-middle-income countries. The date on which a country reported its first case(s) to the WHO can be taken as indicator of the date on which the disease began to spread in that country. Given that indicator, it is clear that the temporal order of the introduction and the start of transmission among income-defined categories is exactly the same as the economic ranking of those categories In other words, overall, the disease has been spreading from higher-income countries to lower-income countries.

A second possible explanation for the observed differences in the number of cases by income-defined groups of countries involves differences in the availability of and approaches to SARS-CoV-2 testing, including testing patients with illness of various severities. In this context two indices of testing are often used: daily tests per capita and test-positivity rate (the number of positive tests as a percentage of the total number of tests done). High-income and upper-middle-income countries are more likely to test more people with illness of various severities than are lower-middle-income and low-income countries. Kobia and Gitaka (2020) also attributed the low apparent incidence rate of COVID-19 in Africa to insufficient testing. In large parts of Africa and also in most low-income and middle-income countries elsewhere, shortages of test kits, lack of capacity to implement large-scale testing and contact tracing, and lack of capacity to roll out surveillance testing have been reported (Jaffer Shah et al. 2020; Kobia and Gitaka 2020), all of which can cause the true incidence rate of COVID-19 in those areas to be underestimated.

The test-positivity rate can be used to assess whether enough testing is being done. High test-positivity rates can indicate that only the sickest patients are being tested, which may imply that more people should be tested. The WHO has issued guidance stating that the positivity rate should be below 5% for at least 14 days before social-distancing measures are relaxed. The actual percentage varied both within and among income-defined groups. As shown on the OWID website, testing was limited in lower-middle-income countries and possibly also in low-income countries, which probably resulted in underestimation of incidence rates (OWID data 2020b).

Third, the income-related differences might also have been caused in part by differences in the timing, adherence to, and/or implementation of preventive and community and public-health measures among the countries. Such measures include frequent handwashing with soap and water, use of alcohol-based hand rub, wearing masks, social distancing, case detection, isolation, contact tracing, and quarantine of exposed persons, closure of schools, public libraries, cinemas, and clubs, and suspension of religious and social gatherings and events, etc. (Baharuddin 2020; CDC COVID-19 response team 2020; Khanna et al. 2020; Lee at al. 2020; Qiu et al. 2020). Large-scale meta-analyses and reviews are required to quantify the impacts of those and other measures on the incidence rate of COVID-19 in different countries.

### Income-related differences in COVID-19 case-fatality rate and death rate per million population

Also noteworthy are the differences in the COVID-19 case-fatality rate, in the daily reported numbers of COVID-19 cases, and in the numbers of deaths per million population, by income-defined group. Overall, higher-income countries were more severely affected in the time period under study. One contributing factor could be longer life expectancy, i.e. the presence of proportionally more elderly people. The spread of COVID-19 in nursing homes for elderly people has been documented (Bedford et al. 2020;Davidson and Szanton 2020;Kemenesi et al. 2020) and older populations have higher prevalences of chronic medical conditions: hypertension, diabetes mellitus, coronary artery disease, chronic obstructive pulmonary diseases, etc. (Suzman and Beard 2015; Zhao et al. 2018). Elderly patients are more likely to have serious and sometimes-fatal medical complications from COVID-19 (Huang et al. 2020). Another possible contributor to the apparent income-related differences is under-reporting of cases and of deaths in lower-income countries due to insufficient surveillance. Nonetheless, the most likely explanation for the bulk of the difference between income-defined groups may still be the relatively late establishment of transmission of the virus in lower-income countries.

### Opposite trends in higher-income and lower-income countries

Another main finding of this study is the fact that the COVID-19 incidence rate decreased in high-income countries from April to May 2020, while at the same time it increased in all other income-defined groups of countries. The impact of the pandemic will be large if the incidence rates already recorded in high-income countries eventually also occur in other countries. Some middle-income and low-income countries have large populations, with many people living in overcrowded conditions, where the virus can easily spread. Moreover, non-pharmaceutical public-health interventions may not be implemented due to a lack of public awareness of their importance. These conditions soon may adversely affect patients, healthcare workers, already-weak health systems, and the economies of those countries (Bong et al. 2020; Jaffer Shah et al. 2000). Already some impact of the pandemic has been observed in middle-income and low-income countries. For example, substantial increases in the numbers deaths have been reported from Ecuador, Brazil, Nigeria, Bangladesh, Pakistan, and Afghanistan (WHO 2020 c). In Guayaquil city of Ecuador, mortuaries were full of corpses of COVID-19 victims and some were even left in the street (The Guardian 2020). In the absence of new medical interventions, the result could be more loss of life than has occurred so far in high-income and upper-middle-income countries.

### Limitations

The findings of this study are subject to some limitations. First, the numbers of cases of COVID-19 reported each day are likely to be underestimated, especially in the middle-income and low-income countries. For example, in Afghanistan, COVID-19 cases have been classified as typhoid fever cases and many symptomatic patients did not present themselves to public health services, due to limited testing services, poor quality of medical care, or stigma associated with COVID-19 (Ariana news 2020; CNA news 2020; Sabawoon 2020). Second, the numbers of deaths reported daily may also be underestimated, because of non-follow-up or incomplete follow-up of patients with COVID-19 who died, or because of deaths among people who were infected with SARS-CoV-2 but in whom COVID-19 was not diagnosed.

## Conclusions

In the first five months of the COVID-19 pandemic, the vast majority of the cases and deaths reported worldwide were from high-income and upper-middle-income countries, with relatively high incidence rates and death rates. Between April and May, the numbers of cases and deaths decreased in high-income countries, but they continued to increase in upper-middle-income, lower-middle-income, and low-income countries.

## Data Availability

All data can be provided based on request.

## Funding

None.

## Conflict of interest

The author declares that he has no conflict of interest.

## Ethical approval and informed consent

Not required.

## Acknowledgement

The author is grateful to Joseph Green (retired from the Graduate School of Medicine at the University of Tokyo) for reading an earlier version of this article and providing comments and suggestions.

## Notes

### Author Declarations

IRB approval not required.

